# Paired CSF and plasma metabolomics reveals a robust CSF-centered metabolic phenotype in idiopathic normal-pressure hydrocephalus: an exploratory case–control study

**DOI:** 10.64898/2026.09.21.26363535

**Authors:** David Visentin, Duje Vukas, Zlatko Kolić, Barbara Kolbah, Mario Lovrić, Stribor Marković, Thomas O. Eichmann, Željka Maglica

**Author notes:** Corresponding author: Željka Maglica.

## Abstract

**Background:** Idiopathic normal pressure hydrocephalus (iNPH) is a potentially treatable neurological disorder whose molecular basis remains incompletely understood. Metabolomic studies in iNPH are scarce and have focused largely on comparisons with other neurological disorders or on cerebrospinal fluid (CSF) alone. Consequently, the broader metabolic phenotype of iNPH and the extent to which CSF alterations are reflected in peripheral blood remain unclear. We therefore compared paired CSF and plasma metabolomic profiles from patients with iNPH and controls.

**Methods:** Paired CSF and plasma samples from 10 patients with iNPH and 10 controls were analyzed via hydrophilic interaction liquid chromatography–mass spectrometry. After quality control and deduplication, 134 CSF and 149 plasma metabolites were quantified. Protein-normalized abundances were log2-transformed and analyzed via age-adjusted ordinary least-squares regression as the primary model, with group-only regression and Mann–Whitney U tests used for sensitivity analyses. Significance was defined as Benjamini–Hochberg q < 0.05 and |log2-fold change| ≥ log2(1.5).

**Results:** Age-adjusted analysis revealed 56 altered CSF metabolites in iNPH patients, 34 of which were increased and 22 of which were decreased; 47 were also significantly altered according to the Mann–Whitney U test. The CSF phenotype encompassed energy and carnitine/acylcarnitine metabolism, redox and one-carbon/transsulfuration-associated metabolism, tryptophan–kynurenine metabolism, amino acid and nucleotide turnover, and neuronal- or membrane-associated metabolites. Prominent changes included increased succinate, 3-hydroxybutyrate, oxidized glutathione, cystathionine, kynurenine and kynurenic acid, together with decreased carnitine, short-chain acylcarnitines, methionine and ergothioneine.

In plasma, 27 metabolites met the age-adjusted significance criteria, but only allantoin, N-formylmethionine, 3-hydroxybutyrate, N-acetylmethionine and kynurenic acid were also significant according to the Mann–Whitney U test. Global CSF–plasma effect–size concordance was moderate. The most consistent cross-compartment features were 3-hydroxybutyrate, N-formylmethionine and N-acetylmethionine.

**Conclusions:** iNPH was associated with a broad and comparatively consistent CSF-centered metabolic phenotype, whereas plasma showed a smaller, partial and model-sensitive reflection of these alterations. These findings support altered energy substrate handling, redox and sulfur-associated metabolism, tryptophan–kynurenine metabolism and neuronal or membrane-associated metabolite handling as components of iNPH biology. These exploratory findings require validation in larger, longitudinal cohorts.

## Background

Idiopathic normal pressure hydrocephalus (iNPH) is a progressive, potentially reversible neurological disorder characterized by gait disturbance, cognitive impairment, and urinary incontinence and is typically accompanied by ventricular enlargement despite normal cerebrospinal fluid (CSF) pressure (1). Its prevalence increases with age, affecting approximately 1.3% of adults aged 65 years and older in pooled epidemiological estimates (2) and up to 8.9% of those over 80 years in prospective population-based studies (3). Despite being one of the few potentially treatable causes of dementia, iNPH remains difficult to diagnose and distinguish from other age-related neurodegenerative disorders, particularly Alzheimer’s disease. Current diagnosis relies primarily on clinical assessment, neuroimaging and CSF drainage, whereas reliable molecular biomarkers for routine diagnostics and prediction of treatment response are lacking (1,4).

Although iNPH was first described in the 1960s, its underlying causes and pathogenesis remain incompletely understood (4). Current evidence supports a multifactorial model in which disturbed CSF dynamics, reduced vascular compliance, altered intracranial pulsatility, cerebral hypoperfusion, glymphatic dysfunction, blood–brain barrier disruption, and secondary inflammatory or metabolic injury interact to drive disease development and progression (5,6). In this model, abnormal CSF pulsatility and impaired CSF drainage may promote ventriculomegaly and periventricular tissue stress, whereas vascular stiffening and impaired cerebral perfusion may contribute to hypoxia-sensitive metabolic dysfunction (7). Glymphatic dysfunction may further link altered CSF dynamics and vascular pulsatility to impaired clearance of interstitial metabolites and neurotoxic proteins (8,9). These mechanisms overlap with processes implicated in Alzheimer’s disease, including impaired amyloid-β clearance, neurovascular dysfunction, glymphatic failure and metabolic stress, with iNPH remaining clinically distinct because symptoms can improve after CSF diversion (9). Molecular signatures that capture iNPH-associated metabolic dysfunction may therefore provide insight into disease biology and help distinguish potentially reversible pathology from other causes of dementia.

Metabolomics offers a direct approach to interrogating these disease-relevant processes, particularly in CSF, which is closely linked to CNS metabolism and clearance. However, metabolomic data in iNPH remain limited. Targeted small-molecule CSF studies have reported reductions in amino acids, isobutyrylcarnitine, citric acid, and dehydroascorbic acid, as well as elevated CSF kynurenic acid in NPH patients, implicating amino acid metabolism, mitochondrial function, oxidative stress and kynurenine pathway activity in disease-associated CSF alterations (10,11). In addition, CSF metabolomics in iNPH/NPH has been limited to a targeted CE‒MS comparison of Alzheimer’s disease versus iNPH, which identified glycerate, N-acetylneuraminate, serine, and 2-hydroxybutyrate as discriminators between the two diseases (12). More recently, a preprint reporting on preoperative and postoperative CSF metabolomics has suggested that metabolic states involving one-carbon/redox metabolism, kynurenine signaling and alternative substrate utilization are associated with neurological recovery after shunt surgery in NPH (13).

Plasma metabolomics in iNPH remains poorly characterized, and it is unclear whether disease-associated metabolic alterations are restricted to the central compartment or reflected in peripheral blood; the only prior report comparing iNPH patients with controls is a small targeted serum profiling study reporting elevated serine in NPH (14).

In the present exploratory study, we performed metabolomic profiling of CSF and plasma from iNPH patients and controls to characterize disease-associated metabolic alterations across biological compartments. We aimed to identify compartment-specific molecular signatures and generate hypotheses regarding metabolic pathways associated with iNPH. Together, this approach provides a comparative view of central and peripheral metabolic alterations in iNPH and helps define the biochemical context of this potentially reversible neurological disorder.

## Methods

### Study design and sample processing

This observational cross-sectional study was conducted over a 1-year recruitment period in collaboration with clinicians at the Clinical Hospital Centre Rijeka and General Hospital Pula. Participants were evaluated as part of routine diagnostic assessment for cognitive impairment and suspected dementia. At each site, experienced clinicians assessed patients for iNPH in accordance with current diagnostic guidelines. Patients with a confirmed diagnosis of iNPH were invited to participate in the study. Control participants were recruited from patients scheduled to undergo elective procedures under spinal anesthesia. All eligible participants received information about the study and provided written informed consent for the use of blood and cerebrospinal fluid samples collected during routine clinical care. In patients with iNPH, CSF and peripheral blood samples were collected during diagnostic lumbar tap testing, whereas in control participants, these samples were collected immediately before the elective procedure.

Peripheral blood was collected in K2EDTA tubes and centrifuged at 6000 rpm for 10 min at 4 °C to obtain plasma, which was separated from the cellular components and aliquoted into sterile 1.5–2 mL microcentrifuge tubes. CSF was collected in sterile additive-free tubes and aliquoted into 0.2–1.0 mL fractions depending on the available volume. All samples were processed under aseptic conditions via a biological safety cabinet and stored at −80 °C until metabolomic and microbiome analyses.

### Metabolite extraction

Metabolite extraction and LC–MS analysis were performed at the Core Facility Mass Spectrometry, Medical University of Graz. Plasma and CSF samples were extracted on ice with cold methanol and water containing a panel of isotopically labeled internal standards (alanine-13C2, serine-13C2, taurine-d4, creatinine-d3, tryptophan-d5, lysine-d3, hypoxanthine-d2, and cholic acid-d4), followed by methyl tert-butyl ether (MTBE) liquid–liquid extraction. After centrifugation, the polar lower phase was collected, dried under vacuum, and reconstituted in 70% acetonitrile containing medronic acid; the protein content was determined via a bicinchoninic acid assay for normalization.

### LC–MS analysis

Chromatographic separation was performed with a Vanquish UHPLC+ system fitted with an ACQUITY UPLC BEH amide column (2.1 × 150 mm, 1.7 µm) operated in both positive (23 min gradient) and negative (18 min gradient) ionization modes. Detection was performed on a Q Exactive Focus mass spectrometer equipped with a heated electrospray ionization source in data-dependent acquisition mode. Pooled quality‒control (QC) samples were injected at the start and end of the sequence and after every fifth sample, alongside extraction blanks and an in-house reference–compound mixture. Metabolites were identified at level 1 (accurate m/z < 5 ppm with retention time and MS2 match to synthetic standards) or level 2; only features with < 25% peak area variation across QC samples were retained. The peak areas were blank-subtracted, normalized to internal standards and to protein content, and expressed in arbitrary units.

### Statistical analysis

Metabolite abundances were log2-transformed prior to analysis. Differential abundance between the iNPH and control groups was assessed separately for each metabolite and each biofluid compartment (CSF, plasma) via ordinary least-squares (OLS) regression with disease group and age as predictors; this age-adjusted model was designated the primary analysis. P values for the group term were adjusted for multiple testing within each compartment via the Benjamini–Hochberg false-discovery-rate (FDR) procedure. A metabolite was considered significantly altered if it met both BH-FDR q < 0.05 and an absolute log2-fold change ≥ log2(1.5).

Because age was imbalanced, age-adjusted results were not interpreted as a complete removal of age-related confounding variables but were considered alongside two sensitivity analyses: a group-only OLS model (excluding age as a covariate) and a two-sided Mann–Whitney U test. P values from both sensitivity analyses were adjusted via the same BH-FDR procedure and fold-change threshold as the primary model. Metabolites significant in both the age-adjusted OLS model and the Mann–Whitney U test are marked with an asterisk in Tables 2 and 3. The global sample structure was examined via principal component analysis (PCA). The log2-transformed abundances were first residualized against age via metabolitewise linear models and then z scored by metabolite before PCA was performed on the residualized, scaled matrix. Differences in multivariate structure between groups were tested by permutational multivariate analysis of variance (PERMANOVA; Euclidean distance, 9999 permutations) applied to this same age-residualized, scaled matrix. Heatmaps were generated from age-adjusted significant metabolites, with log2-transformed abundances scaled rowwise (per metabolite) prior to visualization. CSF–plasma concordance was evaluated among metabolites detected in both compartments by comparing age-adjusted log2-fold changes. Spearman correlation was used to summarize effect-size concordance across all shared metabolites and, separately, among the subset significant in both compartments.

**Table 1.** Cohort and sample characteristics.

| Characteristic | iNPH (n = 10) | Control (n = 10) |
| --- | --- | --- |
| Age, years (mean ± SD) | 71.7 ± 8.6 (range 52 – 81) | 59.7 ± 8.0 (range 41 – 70) |
| Sex, female/male | 4/6 | 4/6 |
| Clinical comorbidities (mean) | 1.9 (range 1 – 4) | 1.7 (range 0 – 4) |
| CSF sample volume, μL | 300 | 300 |
| CSF total protein, μg (mean ± SD) | 10.7 ± 2.8 | 19.2 ± 7.4 |
| Plasma total protein, μg (mean ± SD) | 2847 ± 198 | 3177 ± 375 |
| CSF metabolites quantified, n | 134 | 134 |
| Plasma metabolites quantified, n | 149 | 149 |

Differential-abundance testing (OLS regression, Mann–Whitney U tests) was performed in Python (v_3.12.13.) using pandas (v_2.2.2), SciPy (v_1.16.3) (15), and statsmodels (v_0.14.6). PCA and PERMANOVA (16) were performed in R (v_4.4.1) (17) via vegan (v_2.7.3) (18); figures were generated in R via ggplot2 (v_4.0.3) (19), ggrepel (v_0.9.8), patchwork (v_1.3.2) and pheatmap (v_1.0.13).

## Results

### Study cohort

Twenty participants were included in this preliminary study: 10 patients with iNPH and 10 controls, each providing paired CSF and plasma samples. The sample and dataset characteristics are summarized in Table 1. The groups were partially age-matched (71.7 ± 8.6 vs 59.7 ± 8.0 years; Mann‒Whitney U p = 0.004), with age distributions overlapping between 52 and 70 years and 3 of 10 patients falling inside the control age range. The sex distribution did not differ between the groups (4 females and 6 males in each group; Fisher’s exact test, p = 1.000). The total CSF protein concentration recovered per sample was lower in the iNPH group (10.7 ± 2.8 µg) than in the control group (19.2 ± 7.4 µg), whereas the plasma protein concentration was comparable between the groups. A total of 134 metabolites were quantified in CSF, and 149 were quantified in plasma after deduplication. The comorbidity burden was broadly similar between the groups, averaging approximately 1.9 clinical comorbidities per iNPH participant and 1.7 per control participant when a routine remote surgical history was excluded. Diabetes was more common in the iNPH group (4/10 versus 0/10), whereas arterial hypertension was more common among the controls (5/10 versus 2/10). Other recurrent conditions included cerebrovascular or vascular history; hyperlipidemia; hypothyroidism; and individual neurological, autoimmune, hematologic, or intracranial history.

### CSF metabolome

The CSF metabolome of patients with iNPH was extensively remodeled relative to that of control patients. Among the 134 quantified CSF metabolites, 56 differed significantly between groups after age adjustment (34 increased, 22 decreased; age-adjusted OLS, BH-FDR q < 0.05 and |log2 FC| ≥ log2(1.5); Table 2). Forty-seven of these 56 metabolites were also significant according to the Mann–Whitney U test, indicating that most CSF differences were not dependent on the age-adjusted model alone. Age–metabolite relationships for significant CSF features are shown in Supplementary Figure S1 (Additional file 1). Age-residualized PCA revealed significant group separation, as shown by the PERMANOVA test (R2 = 0.19; p < 0.001), primarily along PC1, with PC1 and PC2 explaining 32.1% and 16.1% of the variance, respectively (Figure 1A). Consistent with this global separation, the volcano plot revealed broad, high-magnitude metabolic differences in both directions (Figure 1B).

**Figure 1.**
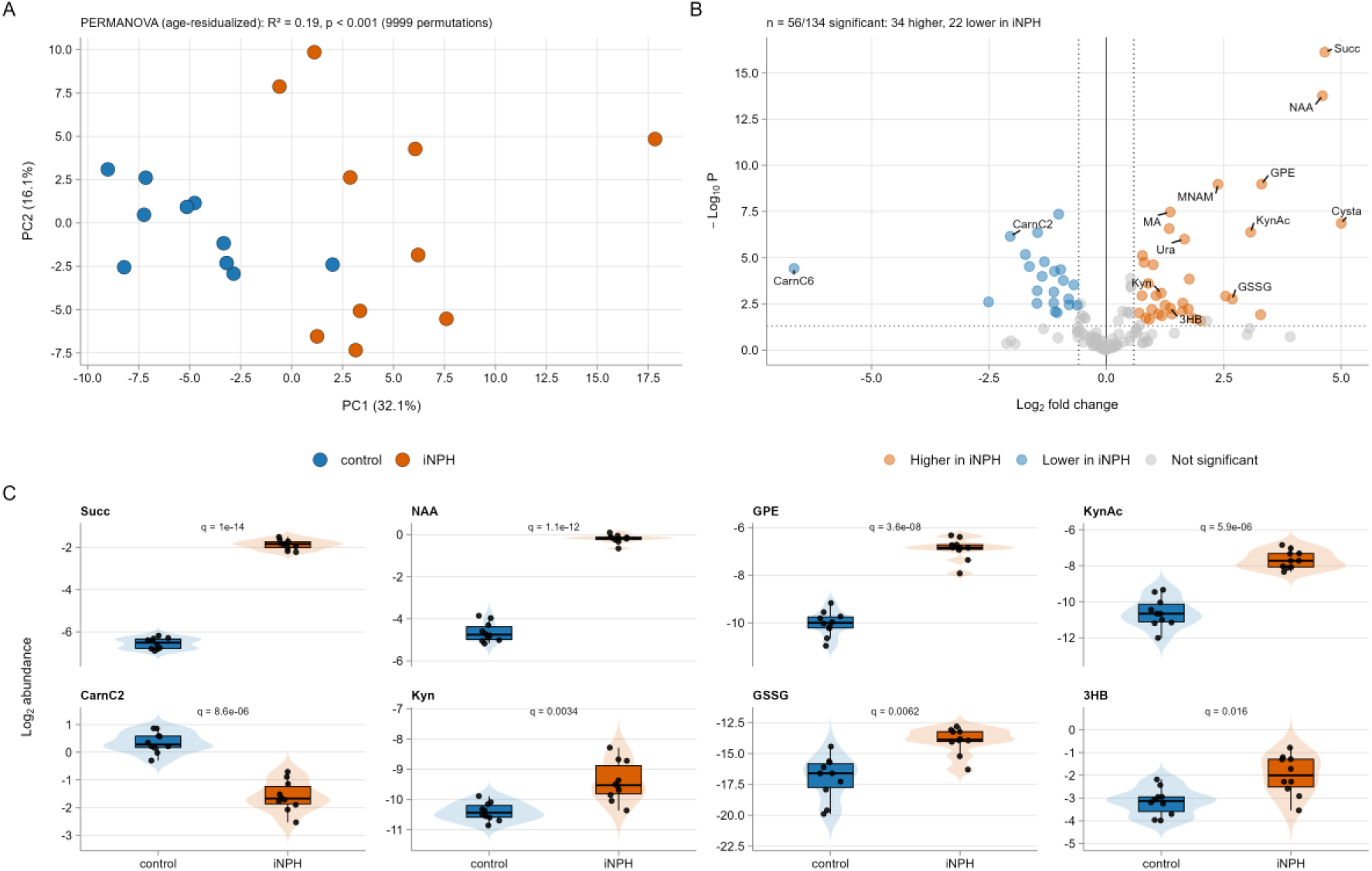
CSF metabolomic phenotype in iNPH patients. (A) Age-residualized PCA of CSF metabolomic profiles with the results of the PERMANOVA test (9999 permutations). (B) Volcano plot of age-adjusted differential abundance. The dotted vertical lines indicate |log2 FC| ≥ log2(1.5), and the dotted horizontal line indicates p = 0.05. (C) Representative headline CSF metabolites showing group-level abundance differences. q values are BH-FDR adjusted values from age-adjusted OLS models. Abbreviations: 3HB, 3-hydroxybutyric acid; 4PA, 4-pyridoxic acid; Alla, allantoin; InSul, indoxyl sulfate; KynAc, kynurenic acid; NAAA, N-amidinoaspartic acid; NAMet, N-acetylmethionine; NFM, N-formylmethionine; SAH, S-adenosylhomocysteine.

The largest-magnitude CSF changes involved mitochondrial, energy and carnitine-associated metabolism. Succinate was the most significantly altered metabolite, with an increase in iNPH CSF (log2 FC = 4.65, q = 1.02 × 10^-14). 3-hydroxybutyric acid was also elevated (log2 FC = 1.36), which is consistent with altered energy-related metabolism (Table 2; Figure 1C). In contrast, free carnitine was decreased (log2 FC = -0.91), alongside a coordinated reduction in acylcarnitines, including acetylcarnitine C2 (-2.04), propionylcarnitine C3 (-1.47), butyrylcarnitine C4 (-1.12), and hexanoylcarnitine C6 (-6.65). Hexanoylcarnitine represented the largest magnitude decrease in the dataset.

Markers of oxidative stress, sulfur metabolism, and methylation-associated pathways were also strongly altered. Among the increased metabolites, cystathionine presented the greatest positive log2-fold change (log2 FC = 5.00), accompanied by increased levels of oxidized glutathione (2.68), 4-pyridoxic acid (2.54), S-adenosylmethionine (0.70), N-acetylmethionine (1.25), methylthioadenosine (1.00), and methylnicotinamide (2.37) (Table 2). In parallel, methionine (- 1.46) and ergothioneine (-2.51) were reduced. Together, these changes are consistent with altered redox, transsulfuration, and one-carbon-associated metabolism in iNPH CSF.

Tryptophan and kynurenine metabolism were strongly represented. Kynurenine (log2 FC = 1.16), kynurenic acid (3.07), 3-hydroxyanthranilic acid (1.11), and indoxyl sulfate (1.40) were increased, which was consistent with altered tryptophan-associated metabolism (Table 2; Figure 1B, C). Amino acid metabolism was also broadly affected, with decreases in the branched-chain amino acids leucine (-1.32), isoleucine (-1.37), and valine (-1.09), as well as proline (-1.47), asparagine (-1.01), threonine (-0.62), and phenylalanine (-0.69). In contrast, the contents of serine (0.80), glutamic acid (1.63), anserine (1.62), guanidinoacetate (1.76), N-amidinoaspartic acid (1.84), and urea (0.97) increased.

Nucleotide, membrane, and neuronal-associated metabolites were further altered. Inosine (-1.64) and uridine (-1.72) decreased, whereas uracil (1.66), hypoxanthine (0.76), orotic acid (1.18), methyladenosine (1.36), and methylimidazolacetic acid (2.01) increased (Table 2). Several metabolites linked to neuronal or membrane-associated metabolism, including N-acetylaspartate (log2 FC = 4.60), glycerophosphorylethanolamine (3.30), and choline (0.77), were among the features associated with the greatest increase in iNPH (Figure 1C). Individual distributions of all age-adjusted significant CSF metabolites are shown in Supplementary Figure S2 (Additional file 1).

**Table 2.** CSF metabolites significantly altered in iNPH versus controls after age adjustment. Age-adjusted OLS, BH-FDR q < 0.05 and |log2 FC| ≥ log2(1.5); n = 56. A positive log2 FC indicates greater abundance in iNPH. *Also significant according to the Mann–Whitney U test (without adjusting for age).

| Compound | Formula | RT (min) | log2 FC | Direction | q (BH-FDR) |
| --- | --- | --- | --- | --- | --- |
| Succinate | C4H6O4 | 6.97 | 4.65 | increased | 1.02e-14* |
| N-Acetylaspartate | C6H9NO5 | 7.13 | 4.6 | increased | 1.15e-12* |
| Glycerophosphorylethanolamine | C5H14NO6P | 10.41 | 3.3 | increased | 3.59e-08* |
| Methylnicotinamide | C7H8N2O | 4.76 | 2.37 | increased | 3.59e-08* |
| Methyladenosine | C11H15N5O4 | 7.59 | 1.36 | increased | 9.12e-07* |
| Asparagine | C4H8N2O3 | 10.04 | -1.01 | decreased | 9.88e-07* |
| Cystathionine | C7H14N2O4S | 12.34 | 5 | increased | 2.63e-06* |
| 4-Acetamidobutyric acid | C6H11NO3 | 1.66 | 1.34 | increased | 4.49e-06* |
| Kynurenic acid | C10H7NO3 | 2.78 | 3.07 | increased | 5.90e-06* |
| Methionine | C5H11NO2S | 7.29 | -1.46 | decreased | 5.90e-06* |
| Acetylcarnitine (C2) | C9H17NO4 | 6.69 | -2.04 | decreased | 8.58e-06* |
| Uracil | C4H4N2O2 | 1.56 | 1.66 | increased | 1.12e-05* |
| Uridine | C9H12N2O6 | 2.29 | -1.72 | decreased | 6.88e-05* |
| Choline | C5H13NO | 3.6 | 0.77 | increased | 7.28e-05* |
| Leucine | C6H13NO2 | 6.39 | -1.32 | decreased | 1.47e-04* |
| Serine | C3H7NO3 | 9.93 | 0.8 | increased | 1.50e-04* |
| Methylthioadenosine | C11H15N5O3S | 1.55 | 1 | increased | 1.85e-04* |
| Inosine | C10H12N4O5 | 4.3 | -1.64 | decreased | 2.20e-04* |
| Hexanoylcarnitine (C6) | C13H25NO4 | 2.58 | -6.65 | decreased | 2.66e-04* |
| Ethanolamine | C2H7NO | 6.88 | -0.97 | decreased | 2.91e-04* |
| N-Acetylputrescine | C6H14N2O | 6.6 | -1.1 | decreased | 3.41e-04* |
| Isoleucine | C6H13NO2 | 6.81 | -1.37 | decreased | 6.17e-04* |
| Guanidinoacetate | C3H7N3O2 | 9.21 | 1.76 | increased | 7.90e-04* |
| Carnitine | C7H15NO3 | 8.45 | -0.91 | decreased | 9.02e-04* |
| Betaine | C5H11NO2 | 6.79 | 0.89 | increased | 1.27e-03* |
| Phenylalanine | C9H11NO2 | 6.2 | -0.69 | decreased | 1.44E-03 |
| Propionylcarnitine (C3) | C10H19NO4 | 4.84 | -1.47 | decreased | 2.71e-03* |
| Creatinephosphate | C4H10N3O5P | 7.93 | -1.12 | decreased | 3.08e-03* |
| Kynurenine | C10H12N2O3 | 6.37 | 1.16 | increased | 3.42e-03* |
| N-Acetylspermidine | C9H21N3O | 10.54 | 1.06 | increased | 4.40e-03* |
| Hypoxanthine | C5H4N4O | 2.73 | 0.76 | increased | 4.40e-03* |
| 4-Pyridoxic acid | C8H9NO4 | 1.33 | 2.54 | increased | 4.54e-03* |
| Glutathione oxidized | C20H32N6O12S2 | 13.57 | 2.68 | increased | 6.16e-03* |
| Ribitol | C5H12O5 | 3.81 | -0.8 | decreased | 6.16e-03* |
| Ergothioneine | C9H15N3O2S | 8.66 | -2.51 | decreased | 8.62e-03* |
| Butyrylcarnitine (C4) | C11H21NO4 | 3.96 | -1.12 | decreased | 9.48e-03* |
| Glutamic acid | C5H9NO4 | 10.27 | 1.63 | increased | 9.48e-03* |
| Proline | C5H9NO2 | 7.89 | -1.47 | decreased | 9.57e-03* |
| Glutamylphenylalanine | C14H18N2O5 | 9.41 | -0.79 | decreased | 1.10E-02 |
| N-Acetylmethionine | C7H13NO3S | 3 | 1.25 | increased | 1.10e-02* |
| Threonine | C4H9NO3 | 9.35 | -0.62 | decreased | 1.10E-02 |
| 3-Hydroxybutyric acid | C4H8O3 | 4.2 | 1.36 | increased | 1.56e-02* |
| Thiamine | C12H16N4OS | 7.21 | 1.74 | increased | 1.71e-02* |
| Urea | CH4N2O | 2.04 | 0.97 | increased | 1.74E-02 |
| Anserine | C10H16N4O3 | 7.45 | 1.62 | increased | 2.10e-02* |
| Valine | C5H11NO2 | 7.8 | -1.09 | decreased | 2.15e-02* |
| N-Acetylarginine | C8H16N4O3 | 9.32 | -1.05 | decreased | 2.33e-02* |
| S-Adenosylmethionine | C15H22N6O5S | 12.11 | 0.7 | increased | 2.39e-02* |
| Indoxyl sulfate | C8H7NO4S | 1 | 1.4 | increased | 2.55E-02 |
| 3-Hydroxyanthranilic acid | C7H7NO3 | 1.75 | 1.11 | increased | 2.55E-02 |
| N-Acetylphenylalanine | C11H13NO3 | 2.32 | 3.28 | increased | 2.76e-02* |
| Orotic acid | C5H4N2O4 | 4.33 | 1.18 | increased | 3.00e-02* |
| N-Formylmethionine | C6H11NO3S | 3.07 | 0.84 | increased | 3.88e-02* |
| N-Amidinoaspartic acid | C5H9N3O4 | 10.3 | 1.84 | increased | 3.96E-02 |
| Glycerate | C3H6O4 | 5.55 | 0.91 | increased | 4.36E-02 |
| Methylimidazolacetic acid | C6H8N2O2 | 8.86 | 2.01 | increased | 4.99E-02 |

A row-scaled heatmap of the top age-adjusted CSF metabolites revealed coordinated group-associated abundance patterns across samples rather than isolated single-metabolite differences (Figure 2). Metabolites increased in iNPH patients, and those reduced in iNPH patients formed broad opposing abundance patterns across the cohort, supporting a structured CSF metabolic phenotype.

**Figure 2.**
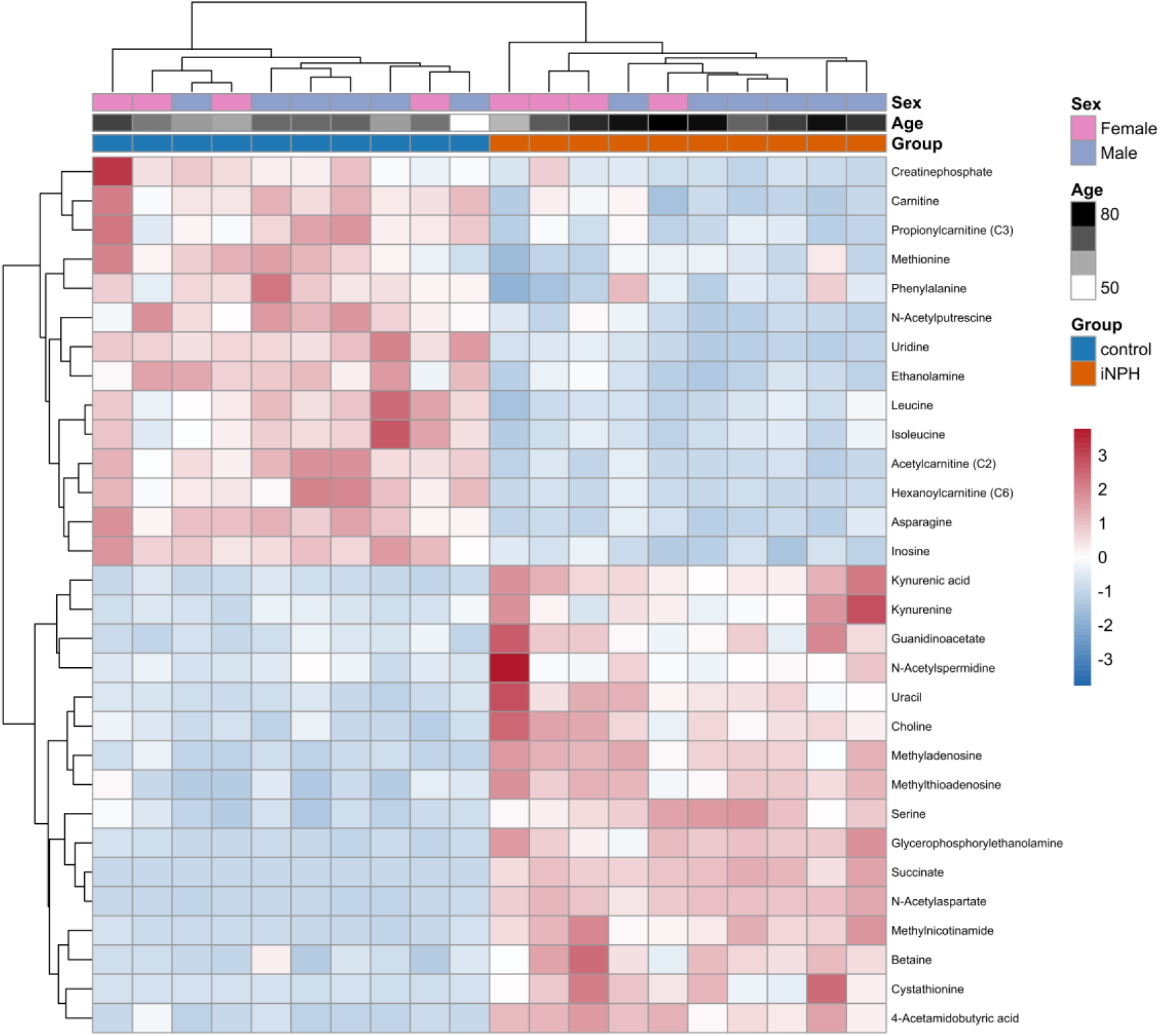
CSF heatmap of the top 30 significant metabolites. Row-scaled heatmap of the top age-adjusted significant CSF metabolites, with sample annotations for group, age, and sex. Metabolites are clustered by abundance pattern across samples.

Taken together, these data indicate a distinct CSF-centered metabolic phenotype in iNPH, characterized by coordinated changes in energy metabolism, carnitine and acylcarnitine balance, oxidative stress and sulfur metabolism, tryptophan‒kynurenine metabolism, amino acid turnover, nucleotide metabolism, and neuronal or membrane-associated metabolites. The persistence of most age-adjusted CSF findings in the Mann–Whitney U test supports the stability of the CSF signal, although residual confounding related to age imbalance cannot be fully excluded.

### Plasma metabolome

In plasma, the disorder-associated metabolomic signal was weaker and more model sensitive than that in CSF. Among the 149 quantified plasma metabolites, 27 differed significantly between groups after age adjustment (20 increased, 7 decreased; age-adjusted OLS, BH-FDR q < 0.05 and |log2 FC| ≥ log2(1.5); Table 3). However, only five of these age-adjusted plasma metabolites were also significant according to the Mann–Whitney U test: allantoin, N-formylmethionine, 3-hydroxybutyric acid, N-acetylmethionine, and kynurenic acid. Thus, the age-adjusted plasma results identified a broader set of candidate metabolites, but most should be interpreted as exploratory and model-sensitive rather than as robust group differences. Regardless, age-residualized PCA revealed significant group separation in plasma, as shown by the PERMANOVA test (R2 = 0.12; p = 0.002), with PC1 and PC2 explaining 31.4% and 10.6% of the variance, respectively (Figure 3A). Unlike the clearer CSF separation, the plasma samples overlapped near the center of the ordination, and there was greater heterogeneity among the iNPH samples. Similarly, the volcano plot revealed a smaller and less symmetric disease-associated signal than that in the CSF (Figure 3B). Individual distributions of all age-adjusted significant plasma metabolites are shown in Supplementary Figure S3 (Additional file 1).

**Figure 3.**
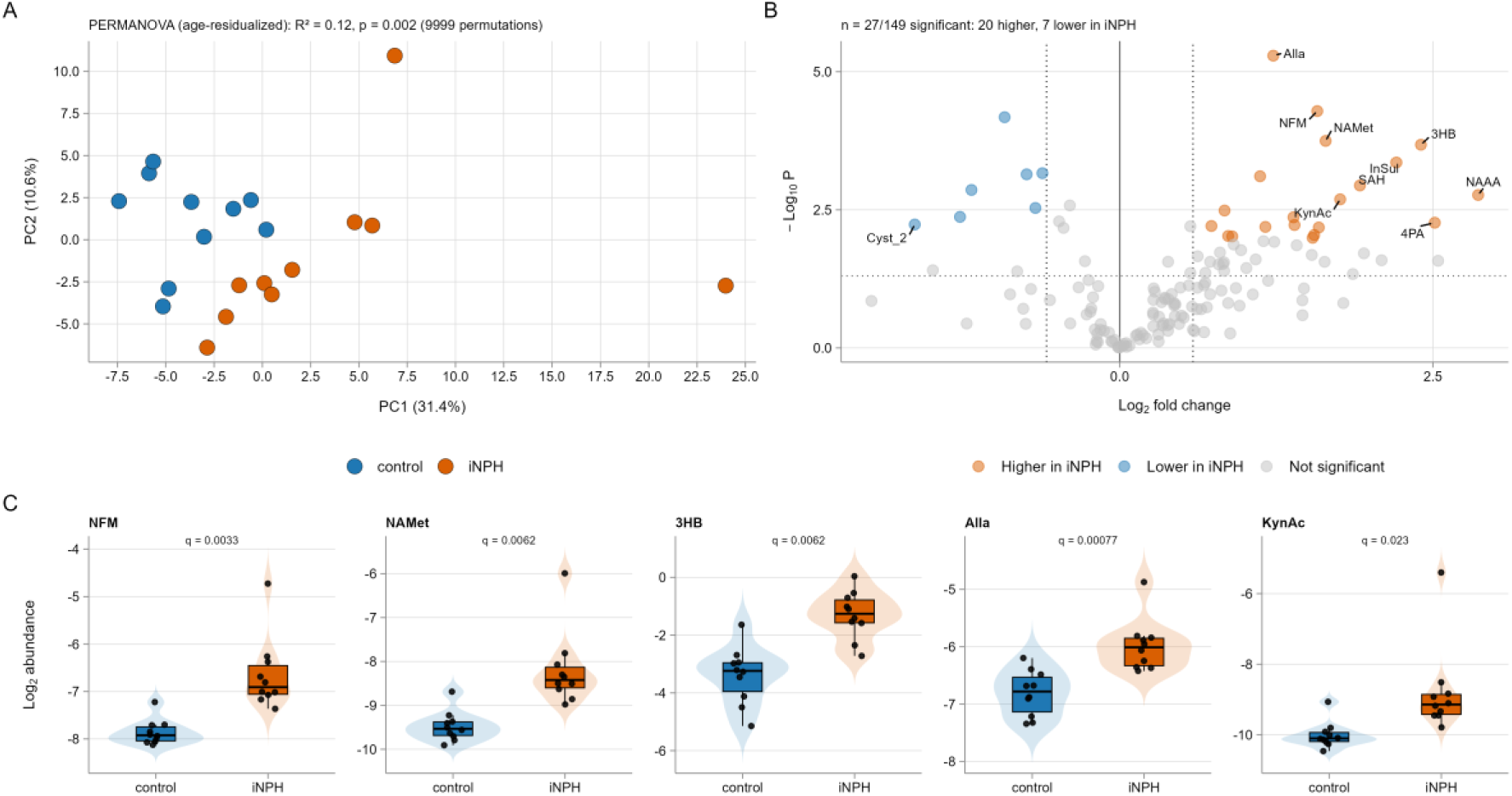
Plasma metabolomic phenotype in iNPH patients. (A) Age-residualized PCA of plasma metabolomic profiles with the results of the PERMANOVA test (9999 permutations). (B) Volcano plot of age-adjusted differential abundance. The dotted vertical lines indicate |log2 FC| ≥ log2(1.5), and the dotted horizontal line indicates p = 0.05. (C) Representative plasma metabolites showing group-level abundance differences. q values are BH-FDR adjusted values from age-adjusted OLS models. Abbreviations: 3HB, 3-hydroxybutyric acid; 4PA, 4-pyridoxic acid; Alla, allantoin; Cyst_2, cystine (isoform 2); InSul, indoxyl sulfate; KynAc, kynurenic acid; NAAA, N-amidinoaspartic acid; NAMet, N-acetylmethionine; NFM, N-formylmethionine; SAH, S-adenosylhomocysteine.

The five metabolites that remained statistically significant according to both age-adjusted OLS and Mann–Whitney U tests were all increased in iNPH plasma. Allantoin had the strongest statistical signal (log2 FC = 1.23, q = 7.66 × 10^-4), followed by N-formylmethionine (1.58, q = 3.30 × 10^-3), 3-hydroxybutyric acid (2.40, q = 6.23 × 10^-3), N-acetylmethionine (1.64, q = 6.23 × 10^-3), and kynurenic acid (1.76, q = 2.34 × 10^-2) (Table 3; Figure 3C). These metabolites indicate that plasma changes involve energy metabolism, methionine-related metabolism, purine oxidation, and tryptophan-kynurenine metabolism.

The broader age-adjusted plasma set included additional metabolites related to amino acid, nucleotide, redox, and microbial-host cometabolism. Tryptophan was decreased (log2 FC = -0.92), whereas kynurenic acid, 3-hydroxyanthranilic acid (1.16), indoxyl sulfate (2.21), and indolelactic acid (0.87) were increased, suggesting a plasma signal involving tryptophan-associated metabolism. The levels of several amino acids or amino acid-related metabolites, including leucine (-0.74), phenylalanine (-1.18), tyrosine (-0.67), ethanolamine (-0.62), and two cystine features (- 1.27 and -1.64), were lower in the plasma of iNPH patients. In contrast, the levels of multiple modified amino acid or nitrogen-containing metabolites, including N-amidinoaspartic acid (2.86), N-acetylasparagine (1.40), N-acetylspermidine (0.90), N-alpha-acetyllysine (1.55), and N-acetylhistidine (1.54), increased.

Nucleotide and redox-associated metabolites were also represented among the age-adjusted plasma findings. Uracil (log2 FC = 1.12), uridine (0.84), xanthine (0.73), S-adenosylhomocysteine (1.92), pyroglutamic acid/oxo-proline (1.39), 4-acetamidobutyric acid (1.59), and 4-pyridoxic acid (2.51) were increased (Table 3). However, because most of these plasma metabolites were not significant according to the Mann–Whitney U test, they should be viewed as age-adjusted candidate signals requiring validation.

**Table 3.** Plasma metabolites significantly altered in iNPH versus controls after age adjustment. Age-adjusted OLS, BH-FDR q < 0.05 and |log2 FC| ≥ log2(1.5); n = 27. A positive log2 FC indicates greater abundance in iNPH. *Also significant according to the Mann–Whitney U test.

| Compound | Formula | RT (min) | log2 FC | Direction | q (BH-FDR) |
| --- | --- | --- | --- | --- | --- |
| Allantoin | C4H6N4O3 | 2.74 | 1.23 | increased | 7.66e-04* |
| N-Formylmethionine | C6H11NO3S | 3.09 | 1.58 | increased | 3.30e-03* |
| Tryptophan | C11H12N2O2 | 4.22 | -0.92 | decreased | 3.30E-03 |
| 3-Hydroxybutyric acid | C4H8O3 | 4.22 | 2.4 | increased | 6.23e-03* |
| N-Acetylmethionine | C7H13NO3S | 3.03 | 1.64 | increased | 6.23e-03* |
| Indoxyl sulfate | C8H7NO4S | 1 | 2.21 | increased | 1.09E-02 |
| Ethanolamine | C2H7NO | 6.88 | -0.62 | decreased | 1.29E-02 |
| Leucine | C6H13NO2 | 6.36 | -0.74 | decreased | 1.29E-02 |
| Uracil | C4H4N2O2 | 1.55 | 1.12 | increased | 1.29E-02 |
| S-Adenosylhomocysteine | C14H20N6O5S | 10.27 | 1.92 | increased | 1.71E-02 |
| Phenylalanine | C9H11NO2 | 6.16 | -1.18 | decreased | 1.87E-02 |
| N-Amidinoaspartic acid (LEU/ILE ION) | C5H9N3O4 | 10.29 | 2.86 | increased | 2.12E-02 |
| Kynurenic acid | C10H7NO3 | 2.81 | 1.76 | increased | 2.34e-02* |
| Tyrosine | C9H11NO3 | 7.91 | -0.67 | decreased | 2.91E-02 |
| Uridine | C9H12N2O6 | 2.3 | 0.84 | increased | 3.04E-02 |
| Cystine_1 | C6H12N2O4S2 | 12.48 | -1.27 | decreased | 3.58E-02 |
| Pyroglutamic acid/oxo-Proline | C5H7NO3 | 5.52 | 1.39 | increased | 3.58E-02 |
| Xanthine | C5H4N4O2 | 3.15 | 0.73 | increased | 3.71E-02 |
| 3-Hydroxyanthranilic acid | C7H7NO3 | 1.74 | 1.16 | increased | 3.71E-02 |
| 4-Acetamidobutyric acid | C6H11NO3 | 1.66 | 1.59 | increased | 3.71E-02 |
| 4-Pyridoxic acid | C8H9NO4 | 1.32 | 2.51 | increased | 3.71E-02 |
| N-Acetylasparagine | C6H10N2O4 | 5.8 | 1.4 | increased | 3.71E-02 |
| Cystine_2 | C6H12N2O4S2 | 7.88 | -1.64 | decreased | 3.71E-02 |
| Indolelactic acid | C11H11NO3 | 2.06 | 0.87 | increased | 4.76E-02 |
| N-Acetylspermidine | C9H21N3O | 10.52 | 0.9 | increased | 4.76E-02 |
| N-alpha-Acetyllysine | C8H16N2O3 | 9.6 | 1.55 | increased | 4.76E-02 |
| N-Acetylhistidine | C8H11N3O3 | 9.33 | 1.54 | increased | 4.89E-02 |

### Compartment specificity and cross-compartment overlap

A comparison of the CSF and plasma analyses revealed that the iNPH-associated metabolomic signal was strongest and most stable in the CSF. The number of significant CSF metabolites was similar across the Mann–Whitney U test, group-only OLS and age-adjusted OLS models, with 50, 53 and 56 significant metabolites, respectively (Figure 4A). In contrast, the plasma signal was smaller and more model sensitive, with 5 significant metabolites according to the Mann–Whitney U test, 6 metabolites according to the group-only OLS and 27 metabolites after age adjustment (Figure 4A). Across metabolites measured in both compartments, CSF and plasma age-adjusted effect sizes showed only moderate global concordance, with most of the shared metabolites clustering near the origin (Spearman ρ = 0.45; Figure 4B). This effect was stronger when metabolites that were significant in both CSF and plasma were observed (Spearman ρ = 0.82; Figure 4C). This significant subset of 15 metabolites presented concordant increases in both compartments, including 3-hydroxybutyric acid, N-formylmethionine, N-acetylmethionine, allantoin, indoxyl sulfate, 4-pyridoxic acid, kynurenic acid and 3-hydroxyanthranilic acid, as well as concordant decreases in leucine, ethanolamine and phenylalanine. Only uridine showed an opposing log2 change between the compartments.

**Figure 4.**
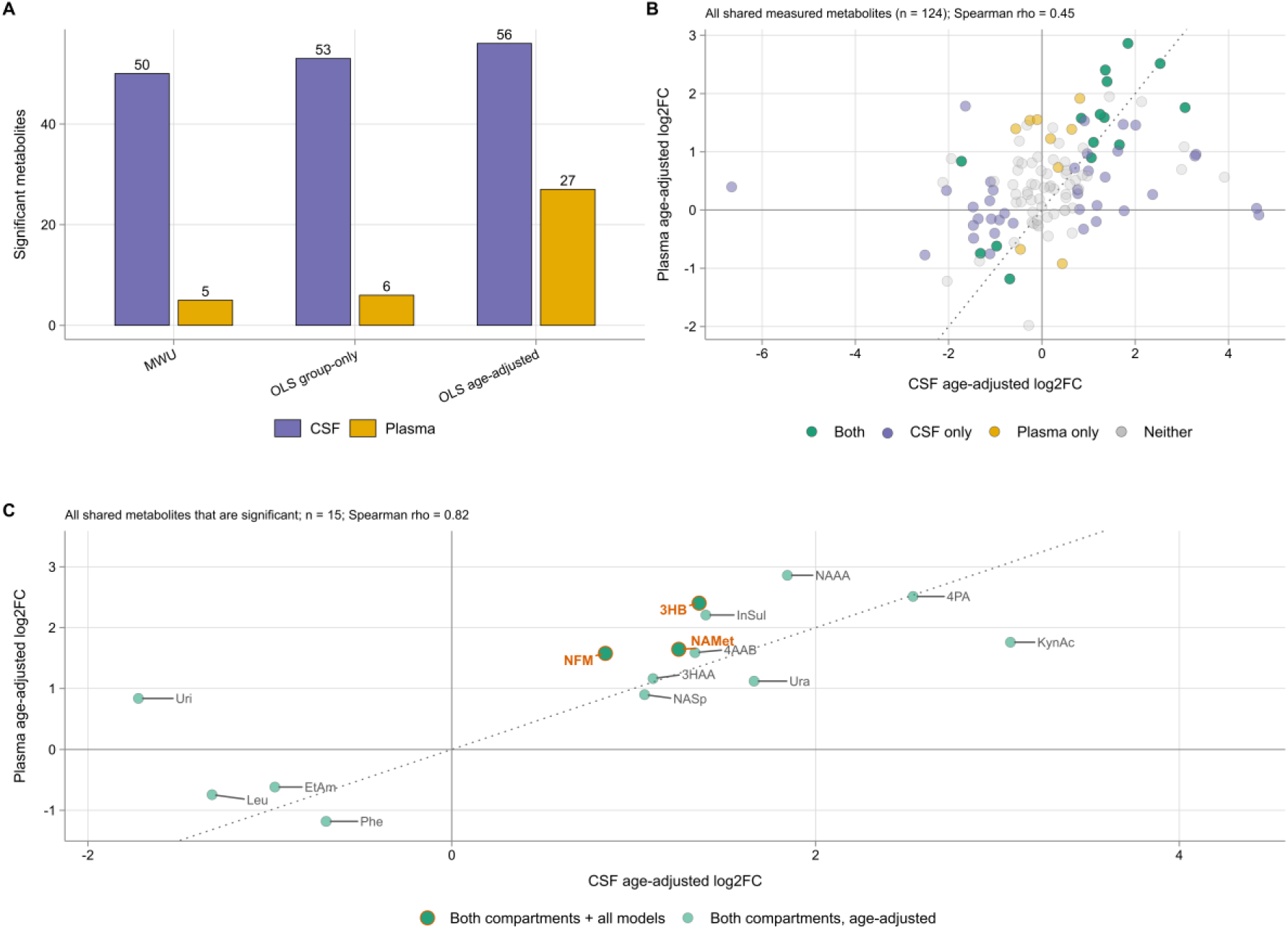
Model sensitivity and CSF–plasma concordance. (A) Significant metabolites in CSF (purple) and plasma (gold) across three models (MWU, OLS group-only, OLS age-adjusted). (B) CSF–plasma log2FC concordance across all shared metabolites (n = 124; Spearman ρ = 0.45), colored by significance in each compartment. (C) Concordance restricted to metabolites significant in both compartments (n = 15; ρ = 0.82); green with orange rims marks metabolites significant and concordant across all CSF and plasma models. Abbreviations: 3HAA, 3-hydroxyanthranilic acid; 3HB, 3-hydroxybutyric acid; 4AAB, 4-acetamidobutyric acid; 4PA, 4-pyridoxic acid; EtAm, ethanolamine; InSul, indoxyl sulfate; KynAc, kynurenic acid; Leu, leucine; NAAA, N-amidinoaspartic acid; NAMet, N-acetylmethionine; NASp, N-acetylspermidine; NFM, N-formylmethionine; Phe, phenylalanine; Ura, uracil; Uri, uridine.

Among the shared metabolites, 3-hydroxybutyric acid, N-formylmethionine and N-acetylmethionine were the most consistent cross-compartment features, remaining significant across all models specified in both CSF and plasma (Figure 4C). These metabolites therefore represent the clearest plasma echo of the stronger CSF-centered phenotype. Overall, these analyses indicate that iNPH is characterized by a broad, robust CSF metabolic phenotype and a smaller plasma component.

## Discussion

In this exploratory paired CSF–plasma metabolomics study, iNPH was associated with a pronounced CSF-centered metabolic phenotype. The primary age-adjusted analysis revealed 56 altered CSF metabolites, 47 of which were also significant according to the Mann–Whitney U test. In contrast, 27 plasma metabolites met the age-adjusted criteria, but only five were supported by the nonparametric analysis. The CSF findings were therefore substantially more stable, whereas the broader plasma profile was weaker and more age sensitive.

The CSF alterations involved several interconnected metabolic domains, including energy and carnitine metabolism, redox and one-carbon/transsulfuration-associated metabolism, tryptophan–kynurenine metabolism, amino acid and nucleotide turnover, and neuronal- or membrane-associated metabolites. A predominant CSF signal is biologically sensible in the context of iNPH, given the central involvement of disturbed CSF dynamics, ventricular enlargement, periventricular tissue stress and altered exchange or clearance at brain–CSF interfaces (5,6,9).

### Energy and mitochondrial substrate metabolism

Energy metabolism is a major component of the iNPH-associated CSF signature. Free carnitine and several short-chain acylcarnitines are reduced together with creatine phosphate, whereas succinate and 3-hydroxybutyrate are increased. This pattern extends previous targeted observations of altered acylcarnitines in iNPH and is consistent with disturbed carnitine/acylcarnitine balance and mitochondrial substrate handling (10,20) and is compatible with reported mitochondrial abnormalities in iNPH brain tissue (21). These changes do not, however, converge on a single mechanism: reduced acylcarnitines do not by themselves demonstrate impaired β-oxidation, and increased 3-hydroxybutyrate may reflect altered production, delivery, utilization or clearance rather than one causal process. Nonetheless, the combined pattern supports altered oxidative and alternative-substrate handling as a genuine feature of the iNPH CSF phenotype.

### Redox and one-carbon/transsulfuration metabolism

**The** iNPH CSF also presented a coordinated redox and sulfur metabolism signature spanning glutathione, the methionine/S-adenosylmethionine (SAM) cycle, and transsulfuration. Oxidized glutathione (GSSG) was increased, whereas methionine and ergothioneine were reduced, a pattern consistent with the elevated oxidative burden reported in other neurodegenerative and neuroinflammatory conditions (22,23). Rather than reflecting simple methyl-donor depletion, the accompanying increase in cystathionine, SAM, betaine, methylthioadenosine (MTA) and methyladenosine, together with elevated N-acetylmethionine and N-formylmethionine, which further point to depletion of the bioavailable methionine pool, suggests remodeling of the methionine/SAM cycle, with increased flux through methylation, methionine salvage, polyamine metabolism and transsulfuration. This is mechanistically grounded: methionine is the precursor of SAM, the principal cellular methyl donor, whereas cystathionine is the first committed intermediate of the transsulfuration pathway, which cells favor under oxidative conditions to supply the cysteine needed for glutathione synthesis; cystathionine β-synthase activity is known to be increased during oxidative stress and is required for astrocyte survival (24,25). MTA and methyladenosine, which are generated during SAM-dependent polyamine synthesis and turnover of methylated nucleic acids, respectively, together with elevated N-acetylspermidine and reduced N-acetylputrescine, indicate that the consequences of methionine depletion extend beyond transsulfuration to polyamine metabolism and nucleotide homeostasis. Reduced ergothioneine, a dietary antioxidant proposed to be protective against age-related neurodegeneration (26), together with reduced dehydroascorbic acid, as reported in recent targeted iNPH CSF metabolomics (10), reinforces the impression that antioxidant defenses are under sustained pressure. Impaired CSF molecular clearance in iNPH could contribute to the concentrations of some of these extracellular metabolites, but its specific contribution to the observed redox signature cannot be determined here. Taken together, the combination of increased GSSG, cystathionine, betaine, MTA and methyladenosine alongside reduced methionine and ergothioneine defines a single coherent oxidative stress and one-carbon metabolism signature in iNPH CSF.

### Amino acid dysregulation

Broad amino acid dysregulation was a further consistent feature of the CSF metabolome. Leucine, isoleucine and valine (branched-chain amino acids; BCAAs) are reduced alongside proline, asparagine, ethanolamine and N-acetylarginine, which is consistent with recent targeted metabolomics reporting broad reductions in proline, threonine, histidine, tyrosine and tryptophan in iNPH (10). Prospective epidemiological evidence links low plasma BCAA levels to increased dementia risk (27), suggesting that BCAA depletion may reflect a metabolic vulnerability shared across conditions associated with cognitive decline; as BCAAs are substrates for cerebral energy metabolism and for glutamate/GABA synthesis via transamination, their depletion may reflect reduced availability for neuronal energetic and synthetic demands, which is consistent with the mitochondrial substrate insufficiency noted above. Among the elevated amino acids, elevated serine stands out, as it echoes prior serum metabolomics in NPH (14). The concurrent depletion of asparagine alongside increased serine and glutamate may reflect altered flux through the aspartate–asparagine–glutamate axis.

### Kynurenine pathway activation

This amino acid signature extends into the kynurenine pathway, the dominant route of tryptophan catabolism in the CNS: both kynurenine and kynurenic acid (KYNA) are elevated, indicating pathway activation and altered neuroimmune-associated metabolic signaling. Elevated CSF KYNA has previously been reported in NPH/iNPH (11), and the concurrent increase in kynurenine extends this observation to an upstream pathway intermediate. Notably, the level of 4-pyridoxic acid, a vitamin B6 catabolite, also increased. This finding is of potential interest given recent evidence linking vitamin B6-dependent metabolism to kynurenine pathway dysregulation in Parkinson’s disease (28). The co-occurrence of increased 4-pyridoxic acid, kynurenine and kynurenic acid therefore raises the possibility of altered B6-associated kynurenine metabolism in iNPH, but this hypothesis requires targeted assessment of vitamin B6 status, pathway ratios and relevant enzyme activity.

### Nucleotide turnover and membrane/neuronal remodeling

Significant alterations in purine and pyrimidine metabolism were also detected. The nucleosides inosine and uridine were reduced, whereas hypoxanthine, uracil and orotic acid were increased. Nucleotide metabolism is closely linked to the cellular energy and redox state, providing biological coherence with the broader energy-related CSF signature observed in this study (29). Nucleotide-related alterations have previously been described in hydrocephalus CSF, although largely in pediatric or acquired hydrocephalus settings (30). Impaired molecular clearance is a plausible contributor to iNPH, given evidence of delayed CSF tracer clearance and abnormal CSF redistribution (31). However, reduced CSF turnover alone would not explain the concurrent reduction in nucleosides and increase in bases. The observed pattern may instead reflect an altered balance between nucleotide degradation, recycling, transport and clearance. These findings provide evidence that nucleotide disturbance contributes to adult iNPH and support purine and pyrimidine metabolism as part of the broader CSF-centered phenotype, whereas targeted measurement of nucleotides, nucleosides and relevant enzymatic steps is needed to define the underlying mechanism.

### Neuronal and membrane metabolism

Increased N-acetylaspartate (NAA), choline and glycerophosphorylethanolamine (GPE) levels indicate altered neuronal and membrane-associated metabolite handling in iNPH CSF. GPE and choline are closely related to phospholipid metabolism, and their elevation is consistent with altered membrane turnover or transport (32). Notably, tissue-based magnetic resonance spectroscopy studies in iNPH have consistently reported reduced intraneuronal NAA concentrations and NAA/creatine ratios (33,34). This is not necessarily contradictory because MRS measures the NAA signal within brain tissue, whereas the present study quantified NAA in CSF. Increased CSF NAA could reflect altered cellular release, transport, utilization or clearance.

### Plasma as a partial peripheral reflection of the CSF phenotype

The plasma findings represent a smaller and less stable counterpart to the CSF phenotype. Twenty-seven plasma metabolites met the primary age-adjusted criteria, but only allantoin, N-formylmethionine, 3-hydroxybutyrate, N-acetylmethionine and kynurenic acid were also significant according to the Mann–Whitney U test. The most consistent cross-compartment features were 3-hydroxybutyrate, N-formylmethionine and N-acetylmethionine, which remained significant across all model specifications in both CSF and plasma and therefore provided the clearest evidence of a peripheral echo of the stronger CSF phenotype. However, the global CSF–plasma effect–size concordance was only moderate, indicating that plasma does not simply reproduce the central metabolic profile. The present data support plasma as a partial peripheral reflection rather than an equivalent surrogate for CSF. The evaluation of any plasma metabolite as a biomarker will require substantially larger, better age-matched cohorts and independent assessment of diagnostic performance.

### Metabolic candidates and pathophysiological model

The present findings identify a focused set of metabolites for targeted validation in future iNPH studies. In CSF, candidates spanning complementary aspects of the observed phenotype include succinate and 3-hydroxybutyrate for energy-related metabolism; free carnitine and representative short-chain acylcarnitines for carnitine-associated substrate handling; oxidized glutathione, cystathionine and methionine for redox and sulfur-associated metabolism; kynurenic acid for tryptophan–kynurenine metabolism; and N-acetylaspartate and glycerophosphorylethanolamine for neuronal- and membrane-associated metabolite handling. This is not intended as a definitive biomarker panel but rather as a biologically informed set of candidates for quantitative replication and assessment against clinical and imaging measures.

In plasma, the strongest candidates for peripheral validation are 3-hydroxybutyrate, N-formylmethionine, N-acetylmethionine, kynurenic acid and allantoin because these metabolites were supported by both the primary age-adjusted analysis and Mann–Whitney U test. Among them, 3-hydroxybutyrate, N-formylmethionine and N-acetylmethionine were the most consistent cross-compartment features.

Together, the findings support a working model (Figure 5) in which reduced CSF turnover and impaired exchange at perivascular and brain–CSF interfaces may have complementary consequences: metabolic stress on periventricular neurons and astrocytes through altered perfusion, mechanical strain and solute exchange, together with slower clearance of extracellular metabolites. The accompanying CSF phenotype is consistent with disturbances in mitochondrial substrate handling and cellular energetics, including reduced carnitine/acylcarnitines and increased succinate and 3-hydroxybutyrate. Concurrent changes in one-carbon- and transsulfuration-associated metabolites and kynurenine pathway intermediates may reflect adaptive responses aimed at maintaining redox homeostasis under this stress, although pathway activation and functional adequacy cannot be inferred from steady-state metabolite concentrations. Altered nucleotide turnover and neuronal- or membrane-associated metabolites may likewise reflect reduced clearance in addition to changes in cellular release or transport. Longitudinal sampling before and after CSF diversion will be needed to determine whether this phenotype is reversible and whether individual metabolites track disease severity or clinical response, a question directly motivated by a recent preoperative CSF metabolomics study reporting that signatures in these same three axes (one-carbon/redox metabolism, kynurenine signaling and alternative substrate utilization) are associated with postoperative neurological recovery in the iNPH (13).

**Figure 5.**
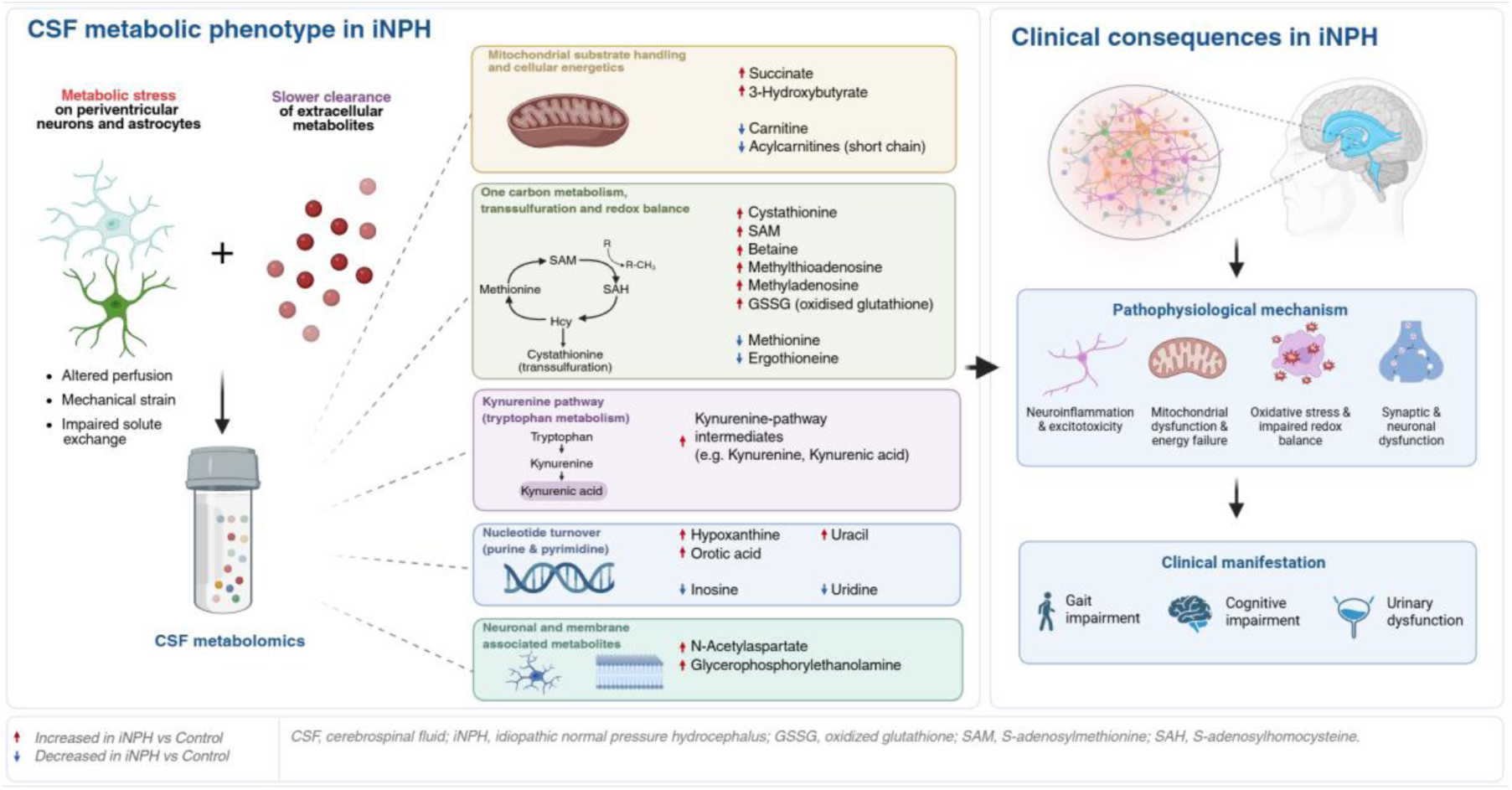
Proposed links between the CSF metabolic phenotype and clinical manifestations of iNPH. Left, altered CSF metabolites are grouped into major metabolic domains associated with periventricular cellular stress and impaired metabolite clearance. These alterations are proposed to contribute to interconnected pathophysiological processes underlying gait, cognitive impairment and urinary impairment. The red and blue arrows indicate increased and decreased metabolites, respectively, in iNPH patients relative to controls. The schematic is hypothesis-generating and does not imply causality. Created in BioRender. Lovric, M. (2026) https://BioRender.com/zw6rkwb

### Limitations and strengths

The main strength of this study is the analysis of CSF and plasma obtained from the same participants, allowing direct comparisons of central and peripheral metabolic patterns. The study also provides broad metabolomic coverage in a condition for which paired-biofluid data remain limited. The use of age-adjusted OLS as the primary differential model, together with group-only OLS and Mann–Whitney U sensitivity analyses, also made model dependence transparent. The marked contrast between the stability of the CSF and plasma results is therefore an informative observation.

The main limitation is the small sample size. The cohort included 10 iNPH patients and 10 controls, limiting statistical power, increasing sensitivity to individual participants and preventing robust adjustment for multiple clinical covariates. The groups were partially age-matched, and only partial age overlap was present. Although the CSF signal remained stable across model specifications and was stronger than the plasma signal was, residual age-related confounding cannot be excluded. Comorbidities are another important limitation. Diabetes was more common in the iNPH group, whereas arterial hypertension was more common among the controls. These conditions may influence energy, redox and vascular metabolism. The small sample size precluded formal adjustment for individual comorbidities, so validation in larger cohorts with more detailed comorbidity matching is needed. Finally, the cross-sectional design limits interpretability and does not address whether the metabolic phenotype changes after CSF diversion or how it is associated with shunt response.

## Conclusion

iNPH is associated with a broad and distinct CSF-centered metabolic phenotype involving energy and carnitine/acylcarnitine balance, redox and one-carbon/transsulfuration-associated metabolism, tryptophan–kynurenine metabolism, amino acid turnover, nucleotide metabolism, and neuronal- or membrane-associated metabolites. Plasma showed a smaller, partial and model-sensitive reflection of this phenotype. These findings provide a biochemical framework for iNPH and identify metabolic features that merit validation in larger, age-matched and longitudinal cohorts, including studies examining their relationship with clinical severity and response to CSF diversion.

## Data Availability

The datasets generated and analyzed during the current study are available from the corresponding author upon reasonable request.

## List of abbreviations

BCAA: Branched-chain amino acid
BH: Benjamini–Hochberg
CNS: Central nervous system
CSF: Cerebrospinal fluid
EDTA: Ethylenediaminetetraacetic acid
FDR: False discovery rate
GABA: γ-Aminobutyric acid
GPE: Glycerophosphorylethanolamine
GSSG: Oxidized glutathione
iNPH: Idiopathic normal pressure hydrocephalus
KYNA: Kynurenic acid
LC–MS: Liquid chromatography–mass spectrometry
MRS: Magnetic resonance spectroscopy
MTA: Methylthioadenosine
MTBE: Methyl tert-butyl ether
NAA: N-Acetylaspartate
NPH: Normal pressure hydrocephalus
OLS: Ordinary least squares
PCA: Principal component analysis
PERMANOVA: Permutational multivariate analysis of variance
QC: Quality control
RT: Retention time
SAH: S-Adenosylhomocysteine
SAM: S-Adenosylmethionine
UHPLC: Ultrahigh-performance liquid chromatography

## Declarations

### Ethics approval and consent to participate

The study was conducted in accordance with the Declaration of Helsinki and applicable national and institutional ethical requirements. It was approved by the Ethics Committee of the Faculty of Biotechnology and Drug Development, University of Rijeka (Class: 644-07/24-01/01; Ref. No. 2170-137-005-01-24-10; 13 December 2024), the Ethics Committee of the Clinical Hospital Centre Rijeka (Class: 003-05/24-01/156; Ref. No. 2170-29-02/1-24-2; 27 December 2024), and the Ethics Committee of Pula General Hospital (Class: 641-01/25-01/01; Ref. No. 2168/01-59-79-112-25-18; 4 March 2025). Written informed consent was obtained from all participants prior to their inclusion in the study.

### Consent for publication

Not applicable.

### Authors’ contributions

Conceptualization, Ž.M., D.Vu. and Z.K.; methodology, D.Vu., Z.K., B.K., D.Vi., M.L., T.O.E., Ž.M. and S.M.; data curation, D.Vu., Z.K., B.K. and D.Vi.; formal analysis, D.Vi., M.L. and T.O.E.; writing-original draft preparation, D.Vi. and M.L.; writing-review and editing, D.Vi., D.Vu., Z.K., S.M., B.K., M.L., T.O.E. and Ž.M.; supervision, Ž.M.; project administration, Ž.M.; funding acquisition, Ž.M. All the authors have read and approved the final manuscript.

## Acknowledgments

Not applicable.

## Additional files

Additional file 1 (.docx). Supplementary figures.

Contains Supplementary Figure S1 showing age–metabolite relationships for significant CSF metabolites, Supplementary Figure S2 showing individual distributions of age-adjusted significant CSF metabolites, and Supplementary Figure S3 showing individual distributions of age-adjusted significant plasma metabolites.

## References

1. Nassar BR, Lippa CF. Idiopathic Normal Pressure Hydrocephalus: A Review for General Practitioners. Gerontol Geriatr Med. 2016 Jan 1;2:2333721416643702. doi:10.1177/2333721416643702

2. Martín-Láez R, Caballero-Arzapalo H, López-Menéndez LÁ, Arango-Lasprilla JC, Vázquez-Barquero A. Epidemiology of Idiopathic Normal Pressure Hydrocephalus: A Systematic Review of the Literature. World Neurosurg. 2015 Dec 1;84(6):2002–9. doi:10.1016/j.wneu.2015.07.005

3. Andersson J, Rosell M, Kockum K, Lilja-Lund O, Söderström L, Laurell K. Prevalence of idiopathic normal pressure hydrocephalus: A prospective, population-based study. PLOS ONE. 2019 May 29;14(5):e0217705. doi:10.1371/journal.pone.0217705

4. Nakajima M, Yamada S, Miyajima M, Ishii K, Kuriyama N, Kazui H, et al. Guidelines for Management of Idiopathic Normal Pressure Hydrocephalus (Third Edition): Endorsed by the Japanese Society of Normal Pressure Hydrocephalus. Neurol Med Chir (Tokyo). 2021;61(2):63–97. doi:10.2176/nmc.st.2020-0292

5. Wang Z, Zhang Y, Hu F, Ding J, Wang X. Pathogenesis and pathophysiology of idiopathic normal pressure hydrocephalus. CNS Neurosci Ther. 2020 Nov 26;26(12):1230–40. doi:10.1111/cns.13526 PubMed PMID: 33242372; PubMed Central PMCID: PMC7702234.

6. Bonney PA, Briggs RG, Wu K, Choi W, Khahera A, Ojogho B, et al. Pathophysiological Mechanisms Underlying Idiopathic Normal Pressure Hydrocephalus: A Review of Recent Insights. Front Aging Neurosci. 2022 Apr 28;14. doi:10.3389/fnagi.2022.866313

7. Qvarlander S, Lundkvist B, Koskinen LOD, Malm J, Eklund A. Pulsatility in CSF dynamics: pathophysiology of idiopathic normal pressure hydrocephalus. J Neurol Neurosurg Psychiatry. 2013 Jul 1;84(7):735–41. doi:10.1136/jnnp-2012-302924 PubMed PMID: 23408066.

8. Costa ML, Casanova-Martinez D, Chen H, Colasurdo M, Kan P. Implications of the glymphatic system in the pathogenesis of normal pressure hydrocephalus: an illustrated scoping review. J Neurosurg. 2025 Mar 28;143(1):135–45. doi:10.3171/2024.12.JNS2420

9. Reeves BC, Karimy JK, Kundishora AJ, Mestre H, Cerci HM, Matouk C, et al. Glymphatic System Impairment in Alzheimer’s Disease and Idiopathic Normal Pressure Hydrocephalus. Trends Mol Med. 2020 Mar 1;26(3):285–95. doi:10.1016/j.molmed.2019.11.008

10. Hofling U, Jakobsson J, Erngren I, Ekman O, Freyhult E, Sreenivasan AP, et al. Targeted CSF metabolomics and conformal prediction improve diagnostic accuracy of normal pressure hydrocephalus. Fluids Barriers CNS. 2026 Feb 7;23(1):34. doi:10.1186/s12987-026-00771-z

11. Kepplinger B, Baran H, Kronsteiner C, Reuss J. Increased Levels of Kynurenic Acid in the Cerebrospinal Fluid in Patients with Hydrocephalus. Neurosignals. 2019 May 4;27(1):1–11.

12. Nagata Y, Hirayama A, Ikeda S, Shirahata A, Shoji F, Maruyama M, et al. Comparative analysis of cerebrospinal fluid metabolites in Alzheimer’s disease and idiopathic normal pressure hydrocephalus in a Japanese cohort. Biomark Res. 2018 Jan 22;6(1):5. doi:10.1186/s40364-018-0119-x

13. Duan L, Tiemeyer ME, Leary OP, Hasbrouck A, Sayied S, Amaral-Nieves N, et al. Cerebrospinal fluid metabolomic profiles associate with neurological recovery after shunt surgery in normal pressure hydrocephalus [Internet]. medRxiv; 2026 [cited 2026 May 20]. p. 2026.03.29.26349660. Available from: https://www.medrxiv.org/content/10.64898/2026.03.29.26349660v1 doi:10.64898/2026.03.29.26349660

14. Orešič M, Anderson G, Mattila I, Manoucheri M, Soininen H, Hyötyläinen T, et al. Targeted Serum Metabolite Profiling Identifies Metabolic Signatures in Patients with Alzheimer’s Disease, Normal Pressure Hydrocephalus and Brain Tumor. Front Neurosci. 2018 Jan 9;11. doi:10.3389/fnins.2017.00747

15. SciPy 1.0: fundamental algorithms for scientific computing in Python | Nature Methods [Internet]. [cited 2026 Jul 30]. Available from: https://www.nature.com/articles/s41592-019-0686-2

16. Anderson MJ. A new method for non-parametric multivariate analysis of variance. Austral Ecol. 2001;26(1):32–46. doi:10.1111/j.1442-9993.2001.01070.pp.x

17. R Core Team. R: A Language and Environment for Statistical Computing [Internet]. Vienna, Austria: R Foundation for Statistical Computing; 2024. Available from: https://www.R-project.org/

18. Dixon P. VEGAN, a package of R functions for community ecology. J Veg Sci. 2003;14(6):927–30. doi:10.1111/j.1654-1103.2003.tb02228.x

19. Wickham H. ggplot2 [Internet]. Cham: Springer International Publishing; 2016 [cited 2026 Jul 30]. (Use R!). Available from: http://link.springer.com/10.1007/978-3-319-24277-4 doi:10.1007/978-3-319-24277-4

20. Reuter SE, Evans AM. Carnitine and Acylcarnitines. Clin Pharmacokinet. 2012 Sep 1;51(9):553–72. doi:10.1007/BF03261931

21. Hasan-Olive MM, Enger R, Hansson HA, Nagelhus EA, Eide PK. Pathological mitochondria in neurons and perivascular astrocytic endfeet of idiopathic normal pressure hydrocephalus patients. Fluids Barriers CNS. 2019 Dec 18;16(1):39. doi:10.1186/s12987-019-0160-7

22. Knudsen E, Tadje J, Coggins C, Venketaraman V. Glutathione and neurodegenerative diseases: immunopharmacological implications. Front Pharmacol. 2026;16:1737199. doi:10.3389/fphar.2025.1737199 PubMed PMID: 41625339; PubMed Central PMCID: PMC12852421.

23. Pham TK, Verber N, Turner MR, Malaspina A, Collins MO, Mead RJ, et al. Glutathione Oxidation in Cerebrospinal Fluid as a Biomarker of Oxidative Stress in Amyotrophic Lateral Sclerosis [Internet]. bioRxiv; 2024 [cited 2026 Jun 23]. p. 2024.07.01.601162. Available from: https://www.biorxiv.org/content/10.1101/2024.07.01.601162v1 doi:10.1101/2024.07.01.601162

24. Niu WN, Yadav PK, Adamec J, Banerjee R. S-Glutathionylation Enhances Human Cystathionine β-Synthase Activity Under Oxidative Stress Conditions. Antioxid Redox Signal. 2015 Feb 10;22(5):350–61. doi:10.1089/ars.2014.5891 PubMed PMID: 24893130; PubMed Central PMCID: PMC4307034.

25. Vitvitsky V, Thomas M, Ghorpade A, Gendelman HE, Banerjee R. A Functional Transsulfuration Pathway in the Brain Links to Glutathione Homeostasis*. J Biol Chem. 2006 Nov 24;281(47):35785–93. doi:10.1074/jbc.M602799200

26. Wu LY, Cheah IK, Chong JR, Chai YL, Tan JY, Hilal S, et al. Low plasma ergothioneine levels are associated with neurodegeneration and cerebrovascular disease in dementia. Free Radic Biol Med. 2021 Dec 1;177:201–11. doi:10.1016/j.freeradbiomed.2021.10.019

27. Tynkkynen J, Chouraki V, van der Lee SJ, Hernesniemi J, Yang Q, Li S, et al. Association of branched-chain amino acids and other circulating metabolites with risk of incident dementia and Alzheimer’s disease: A prospective study in eight cohorts. Alzheimers Dement. 2018;14(6):723–33. doi:10.1016/j.jalz.2018.01.003

28. Wilson EN, Umans J, Swarovski MS, Minhas PS, Mendiola JH, Midttun Ø, et al. Parkinson’s disease is characterized by vitamin B6-dependent inflammatory kynurenine pathway dysfunction. NPJ Park Dis. 2025 Apr 26;11:96. doi:10.1038/s41531-025-00964-7 PubMed PMID: 40287426; PubMed Central PMCID: PMC12033312.

29. Integrated Functions of Cardiac Energetics, Mechanics, and Purine Nucleotide Metabolism - Lopez-Schenk - 2024 - Comprehensive Physiology - Wiley Online Library [Internet]. [cited 2026 Jul 28]. Available from: https://onlinelibrary.wiley.com/doi/10.1002/j.2040-4603.2024.tb00292.x

30. Mueller RA, Rosner MJ, Ghia JN, Powers SK, Kafer ER, Hunt RD. Alterations in cerebrospinal fluid uridine, hypoxanthine, and xanthine in head-injured patients. Cell Mol Neurobiol. 1988 Jun 1;8(2):235–43. doi:10.1007/BF00711249

31. Eide PK, Ringstad G. Delayed clearance of cerebrospinal fluid tracer from entorhinal cortex in idiopathic normal pressure hydrocephalus: A glymphatic magnetic resonance imaging study. J Cereb Blood Flow Metab. 2019 Jul 1;39(7):1355–68. doi:10.1177/0271678X18760974

32. Walter A, Korth U, Hilgert M, Hartmann J, Weichel O, Hilgert M, et al. Glycerophosphocholine is elevated in cerebrospinal fluid of Alzheimer patients. Neurobiol Aging. 2004 Nov 1;25(10):1299–303. doi:10.1016/j.neurobiolaging.2004.02.016

33. Lenfeldt N, Hauksson J, Birgander R, Eklund A, Malm J. IMPROVEMENT AFTER CEREBROSPINAL FLUID DRAINAGE IS RELATED TO LEVELS OF N-ACETYL-ASPARTATE IN IDIOPATHIC NORMAL PRESSURE HYDROCEPHALUS. Neurosurgery. 2008 Jan;62(1):135. doi:10.1227/01.NEU.0000311070.25992.05

34. Lundin F, Tisell A, Leinhard OD, Tullberg M, Wikkelsö C, Lundberg P, et al. Reduced thalamic N-acetylaspartate in idiopathic normal pressure hydrocephalus: a controlled 1H-magnetic resonance spectroscopy study of frontal deep white matter and the thalamus using absolute quantification. J Neurol Neurosurg Psychiatry. 2011 Jul 1;82(7):772–8. doi:10.1136/jnnp.2010.223529 PubMed PMID: 21217158.

