## Additional file 1 for "Paired CSF and plasma metabolomics reveals a robust CSF-centered metabolic phenotype in idiopathic normal-pressure hydrocephalus: an exploratory case–control study"

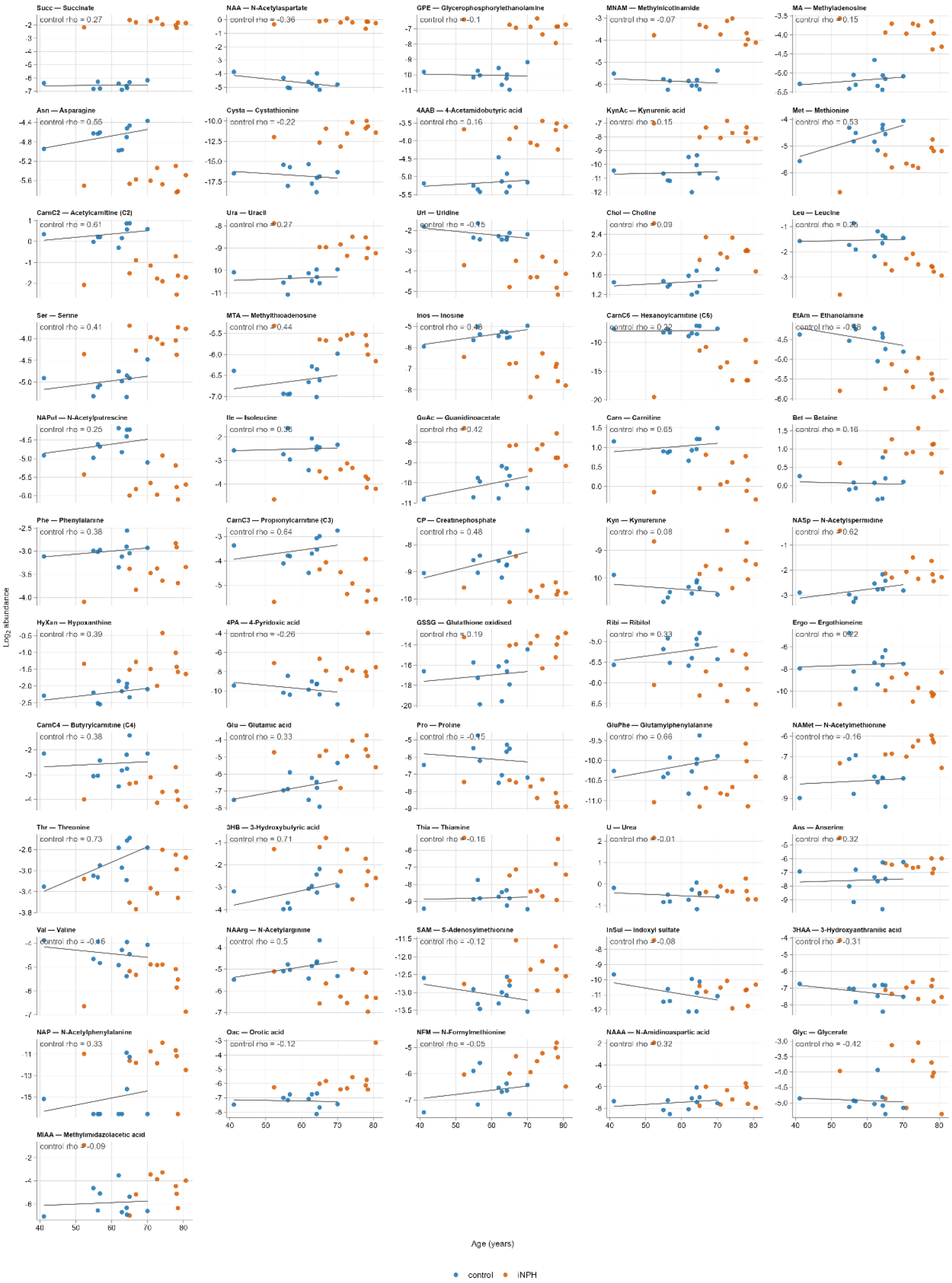


***Supplementary Figure S1. Age–abundance relationships for all age-adjusted significant CSF metabolites.*** *Scatter plots of $\log_2$ abundance versus age for each of the 56 CSF metabolites significant under the age-adjusted OLS model, colored by group (control, iNPH). The fitted line and Spearman correlation ("control rho") are computed within the control group only, to visualize each metabolite's baseline age-dependence independent of disease status. n = 10 iNPH, 10 control.*

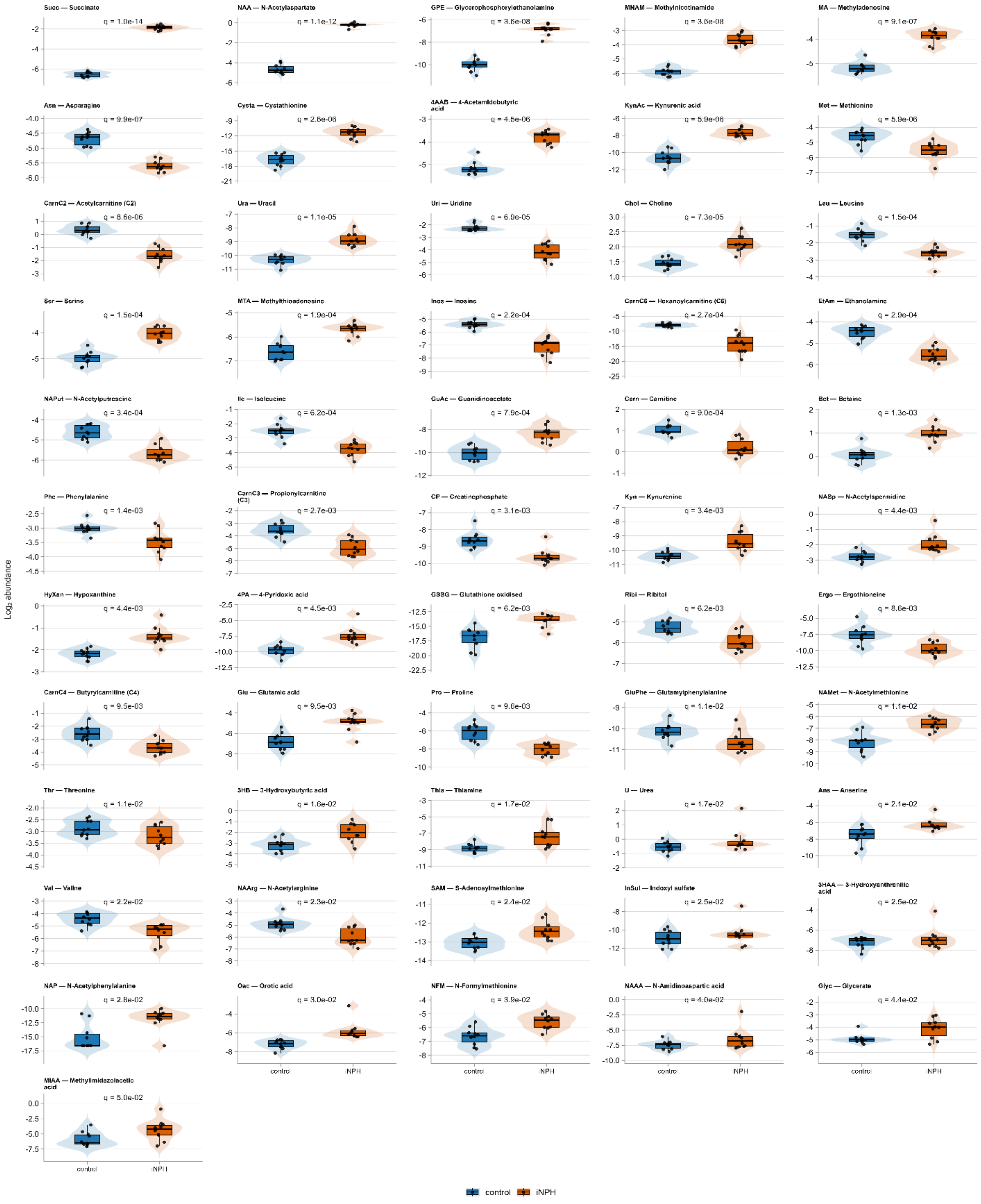


***Supplementary Figure S2. All age-adjusted significant CSF metabolites.*** *Boxplots (median, IQR) with overlaid violin distributions and individual data points for all 56 CSF metabolites significant under the age-adjusted OLS model, ordered by increasing q-value. q-values shown above each panel. n = 10 iNPH, 10 control; groups are age-imbalanced, which is not corrected for in these plots.*

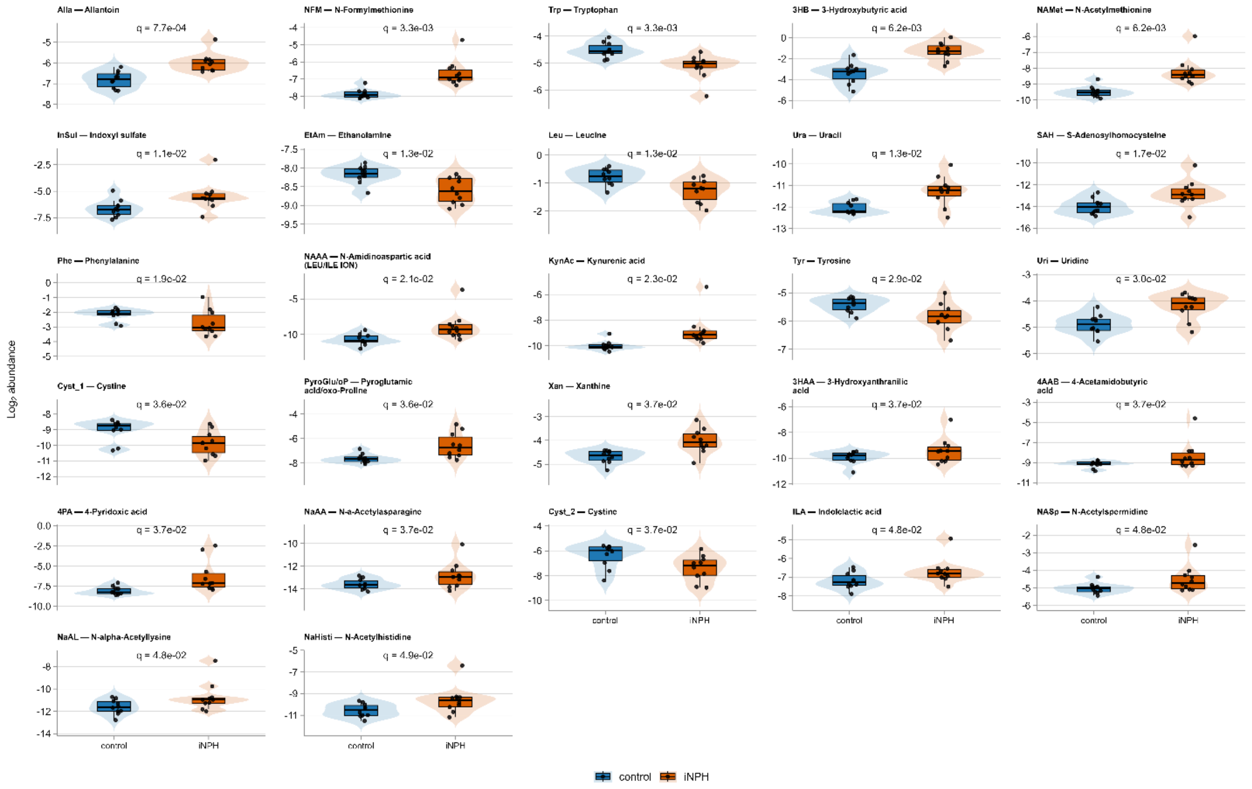


***Supplementary Figure S3. All age-adjusted significant plasma metabolites.*** *Boxplots (median, IQR) with overlaid violin distributions and individual data points for all 27 plasma metabolites significant under the age-adjusted OLS model, ordered by increasing q-value. q-values shown above each panel. n = 10 iNPH, 10 control; groups are age-imbalanced, which is not corrected for in these plots.*
